# Declines in HIV Testing and Diagnoses: A Policy Analysis of the 2019 Title X Federal Regulations on Family Planning Clinics

**DOI:** 10.1101/2025.08.14.25333686

**Authors:** Jennifer Sherwood, Nathan Roberson, Deborah Stenoien, Elise Lankiewicz, Brian Honermann, Greg Millett

## Abstract

In 2019 the Trump administration instituted a federal regulation (hereinafter the Policy) requiring that Title X family planning clinics have financial and physical separation of abortion services from other health services and prohibited abortion referrals. In response, 32% (1,280) of Title X funded sites left the program nationally. Our analysis connects public data from Title X Family Planning Reports (2016-2021) and CDC surveillance reports (2016-2021), to examine the Policy’s impact on: 1) HIV testing at Title X clinics by region, and 2) the proportion of a region’s total HIV diagnoses at Title X clinics. We use mixed multivariable linear regression modeling to examine interactions between high regional-level Policy exposure (defined as >25% Policy-related clinic withdrawal from the program) and pre- and post-Policy implementation for HIV outcomes. Interaction models showed that the Policy resulted in 69,626 fewer HIV tests (95% CI -108,893 to -30,359 P<.001) and a 4% reduction (95% CI -7% to -0%; P =.052) in overall regional HIV diagnoses at Title X clinics in high exposed regions compared to low exposed regions from pre- to post-Policy. Results show notable declines in HIV testing and the proportion of HIV cases diagnosed at Title X sites as a result of Policy implementation. Policies that endanger the Title X family planning network also weaken the U.S. HIV response. Future policies governing the Title X family planning program should consider the full consequences for sexual and reproductive health outcomes, including HIV, in the U.S. before implementation.

**PLAIN LANUGAGE SUMMARY:** The Title X Family Planning Program is a key provider of sexual and reproductive health services in the United States (U.S.), especially for low-income women. It is important to examine any policies governing this safety net program to ensure that we continue to provide these populations with the best care. In 2019 the Trump administration passed a federal rule (hereinafter the Policy) which required that any clinics which receive Title X funding must have “financial and physical separation” of abortion services from other health services and prohibited clinics from making abortion referrals for their patients. About a third (32%) of clinics in the Title X program across the U.S. refused to comply with these restrictions and gave up federal Title X funding. Given the important role of Title X clinics for HIV testing services, we examined what the impact of this mass clinic withdrawal had on HIV testing services and HIV diagnoses in the Title X program. We found that there were significant reductions in both HIV testing and diagnoses in the Title X program related to the Policy. Our results show how policies that reduce providers in the Title X family planning network also weaken the U.S. HIV response. Sound health policy that advances efforts to end the U.S. HIV epidemic will support reproductive health clinics, such as those in the Title X program, as key providers of HIV services.

## INTRODUCTION

The Office of Population Affairs’ Title X Family Planning Program (Title X) is a crucial provider of contraception, sexually transmitted infection (STI) and HIV testing, cancer screenings, and pregnancy counseling in the United States. Federal Title X grants are competitively awarded to state health departments, non-profit organizations, and community health centers with the primary intent of supporting the sexual and reproductive health of low income and uninsured or under-insured Americans.^1^ In March 2019, the Department of Health and Human Services (HHS) under the Trump Administration instituted a regulation (hereinafter “the Policy”) requiring financial and physical separation between Title X funded programs and facilities where abortion is provided.^2^ The 2019 Policy further specified that Title X funded providers were mandated to refer pregnant clients for prenatal care and could not refer clients for abortion services.^3^ In response, 32% (1,280) of the nation’s Title X funded sites refused to comply with the Policy and left the program between June 2019 and August 2021.^4,5^

This mass withdrawal of Title X sites had a significant impact on the program’s family planning service delivery. In 2019, Title X funded clinics served 844,000 fewer family planning users than in 2018.^6^ The 2019 Family Planning Annual Report cites the change in Title X program regulations as a key reason for the reduced capacity and performance of the service network.^6^ In 2020, the effects of the Policy were compounded by the COVID-19 pandemic and Title X funded clinics served 2.4 million fewer family planning users than in 2018.^7^ Of this total reduction, an estimated 63% was attributed to the Policy, while 37% was attributed to the COVID-19 pandemic.^7^ The effects of the Policy on other health services offered at Title X sites, such as HIV testing, have not been widely reported. Examining the effect of the Policy on HIV testing and diagnoses is an important research area given the historical contribution of Title X sites to the U.S. HIV testing infrastructure and the potential for Policy-related disruptions to the U.S. HIV response.^8^

In November of 2021, the Biden Administration finalized a new set of Title X regulations to reverse those of the Trump Administration. As a result, clinics began to re-join the Title X program. By May 2023, the total Title X network had 2% more sites than prior to the Trump era regulations.^58^ However, this progress is fragile. New federal regulations governing the Title X program can be issued at any time and the health impacts of previous drops in service delivery can persist even when the Policy is not in place. This paper examines the impact of the 2019 Policy on HIV testing and diagnoses within the Title X program to inform future discussions on policies governing the Title X family planning program.

## METHODS

Institutional Review Board approval was not required for this secondary analysis using publicly available data. The unit of analysis for this study is U.S. region (as defined by the U.S. Department of Health and Human Services [HHS], available in Appendix 1)^9^ based on data availability.

### Variables and Data Sources

#### Primary exposure

The primary exposure variable is exposure to the Policy by HHS region. Exposure is binary with high exposure regions defined as a net loss of greater than or equal to 25% of the region’s Title X clinics, and low exposure regions defined as a net loss of less than 25% of the region’s Title X clinics from June 2019 to June 2020.^4^ Clinics were considered “lost” if they withdrew from the Title X program during this period and were considered “added” if they newly joined the Title X program in this period. Five regions were coded high exposure and five were coded low exposure (Appendix 1). Data were taken from Health and Human Services Office of Population Affairs’ Title X Directories for 2019 and 2020 collated by the Kaiser Family Foundation.^4^

#### Primary outcomes

This analysis has two primary outcomes: 1) The number of HIV tests delivered at Title X clinics by region, and 2) The proportion of total HIV cases diagnosed at Title X clinics out of all incident HIV diagnoses by region. Data on the number of HIV tests and diagnoses at Title X clinics were taken from the Title X Family Planning Annual Report National Summaries from 2016 to 2021. ^3,6,7,10–12^ This was divided by the total number of regional HIV cases in that year – taken from the Centers for Disease Control (CDC) annual HIV surveillance reports for 2016 to 2021.^13–18^

#### Covariates

Time/year is included as a binary indicator for pre-policy (2016-2018) versus post-policy (2019-2021). Covariates include demographic variables for Title X clients and state-level descriptive variables measured at baseline (2018) aggregated at the region-level. Demographic variables include the proportion of Title X clients by sex (female, male), race (Black, White, and Asian), income (under 101% of federal poverty level, 101%-150%, 151%-200%, and over 201%), and insurance type (uninsured, public, and private) by region. Data on Title X clients come from the Title X Family Planning Annual Report National Summaries 2018.^11^ Region-level descriptive variables, aggregated from state-level data, include; 1) the number of total HIV diagnoses in 2018, from the CDC HIV surveillance 2018 report; ^15^ 2) the proportion of states in region that expanded Medicaid eligibility for people up to 138% of the federal poverty level by 2018, from the 2018 Government Accountability Report on Medicaid expansion,^19^ and; 3) the number of Title X clinics operating by region in 2018, from Title X Family Planning Annual Report 2018 National Summary.^11^ Regional-level covariates were selected to include variables that could influence HIV testing and diagnoses rates at Title X besides the Policy.

### Data Analysis

All data were analyzed in Stata 16.^20^ Statistical significance was set at p<.05.

#### Baseline analysis

All covariates were tested for their association with exposure at baseline (2018). A two-tailed, two-sample t-test for differences was used to assess the difference in client-demographics and region-level descriptive variables among high exposure and low exposure regions. Only covariates significant at baseline (p<.05) were included in modelling given limited power. Likelihood ratio tests were used to assess improved model fit of additional covariates.

#### Mixed effect models

Given the nested nature of our data (regional clusters) with anticipated and known regional variances, we fit mixed linear regression with random intercepts and otherwise fixed regional covariates using maximum likelihood estimation for each outcome (HIV tests per region and proportion of a region’s HIV diagnoses identified at Title X clinics). Models assumed independent covariance with Gaussian errors and observed information matrix standard errors. Regional random intercepts were included with simultaneous estimation on conditional likelihoods. Equations are available in Appendix 2.^9^

A sequential linear mixed-effect modeling approach was used to estimate conditional likelihood on the outcomes. First, we fit null models including random intercepts for regions without covariates. The intraclass correlation coefficient (ICC) was calculated to estimate the proportion of variance attributed to regional random intercepts. Second, we included regional covariates for Policy exposure, pre/post Policy time, and covariates which were significant at baseline including the proportion of Title X clients on public health insurance in 2018, and the proportion of states that expanded Medicaid in 2018.

In the final models, an interaction term was added for exposure by pre/post Policy time as the primary difference-in-difference estimator. The interaction term tests if the outcome differs significantly between Policy time periods by exposure group. Improved fit was evaluated using likelihood ratio tests (LRT) to examine if inclusion of additional parameters was justified.

Pseudo effect sizes were calculated for the interaction terms by taking the difference between the variance explained by the interaction model compared to the regional covariates model as a proportion of total explained variance in the interaction model.

#### Sensitivity Analyses

Authors explored both various model constructions on the outcomes and adjustments to the exposure variable. Model selection is discussed here, and results are discussed, but not shown, below.

Given the limited regional sample size (*n=10*), single-level ordinary least squares (OLS) regression with 9 region binary indicators was considered but given problems with collinearity and region exclusions this approach was dismissed. Single-level OLS with clustered standard errors using the region was also explored. Results were not meaningfully different, and clustered errors can be even more demanding of a large cluster *n*, and so our mixed model is preferred.

Primary exposure was also adjusted in sensitivity analysis at a net loss of clinics greater than or equal to 10%, 15% and 30% compared to our original selection of 25%, and an ordinal coding of the four greatest net loss regions coded as 1; the four smallest net loss coded as 0, and the middle two regions coded as .5.

## RESULTS

### Baseline results and descriptive trends

At baseline (2018), high and low exposure regions were similar on demographic and region-level variables, with two exceptions (Table 1). Clinics in high exposure regions served a higher percentage of people on public insurance (41% vs. 30%, P = .048) and higher levels of Asian clients (55% vs. 22%, P = .007). Among region-level variables, there was a two-tailed, significant difference only in the proportion of states who had expanded Medicaid in high exposure and low exposure regions (97% vs. 58%, P =.028).

**Table 1.** Baseline comparison of high and low exposure regions (2018)

|  | All Regions<br>(n=10) | High Exposure<br>Regions (n=5) | Low Exposure<br>Regions (n=5) | *P* Value <sup>a</sup> |
| --- | --- | --- | --- | --- |
| **Title X Client Demographics** |  |  |  |  |
| Female: Avg. % | 87 | 88 | 87 | .681 |
| Race: Avg. % |  |  |  |  |
| Black | 20 | 17 | 24 | .377 |
| White | 59 | 54 | 64 | .185 |
| Asian | 3 | 5 | 2 | .008* |
| Income level: Avg % |  |  |  |  |
| Under 101% | 62 | 59 | 64 | .309 |
| 101% - 150% | 15 | 16 | 14 | .145 |
| 151% - 200% | 8 | 8 | 7 | .298 |
| Over 201% | 13 | 14 | 12 | .486 |
| Insurance status: Avg. % |  |  |  |  |
| Uninsured | 39 | 33 | 45 | .121 |
| Public | 35 | 41 | 30 | .048* |
| Private | 24 | 24 | 24 | .974 |
| **Regional-Level Variables** |  |  |  |  |
| Total HIV diagnoses:<br>Regional avg. | 3,775 | 3,188 | 4,362 | .603 |
| % States expanded Medicaid | 77 | 97 | 58 | .028* |
| Number of Title X clinics:<br>Regional avg. | 395 | 318 | 473 | .314 |

<sup>a</sup> Two-sample t-test for difference in means
\* significant p<.05

Overall, Title X clinics provided an annual average of 913,903 HIV tests and diagnosed 7.1% of the country’s HIV cases from 2016-2021. Prior to the 2019 Policy, high exposure regions conducted an average of 203,759 more HIV tests (Figure 1) and diagnosed an average of 1.3% more of a region’s total HIV cases (Figure 2) every year. This trend reversed in 2019, with high-exposure regions conducting 67,242 fewer HIV tests and diagnosing 2.8% less of a region’s HIV cases than low exposure regions. These gaps persisted or widened in 2020, with clinics in high exposure regions conducting 183,411 fewer HIV tests and diagnosing 2.0% less of a region’s HIV cases than in low exposure regions.

**Figure 1.**
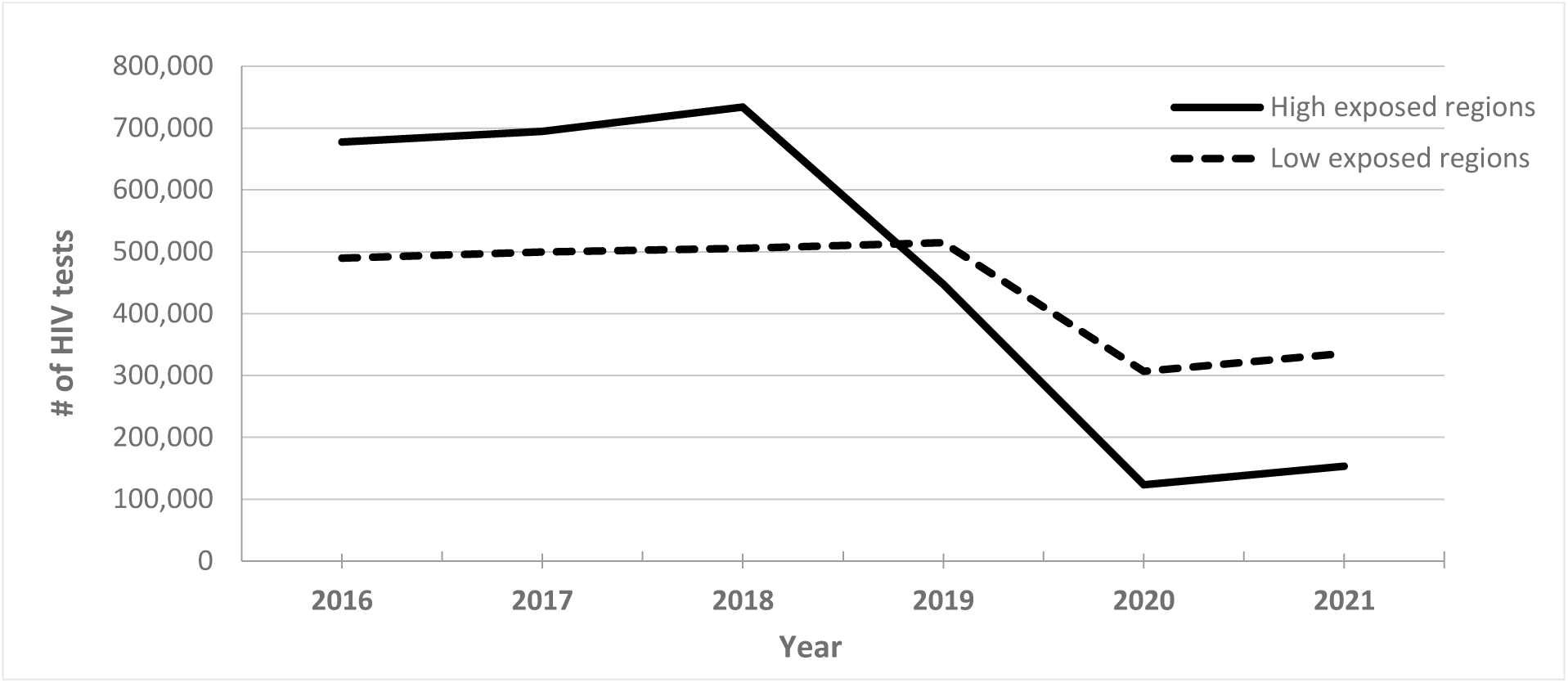
Number of HIV tests provided in high vs. low exposure regions from 2016-2021.

**Figure 2.**
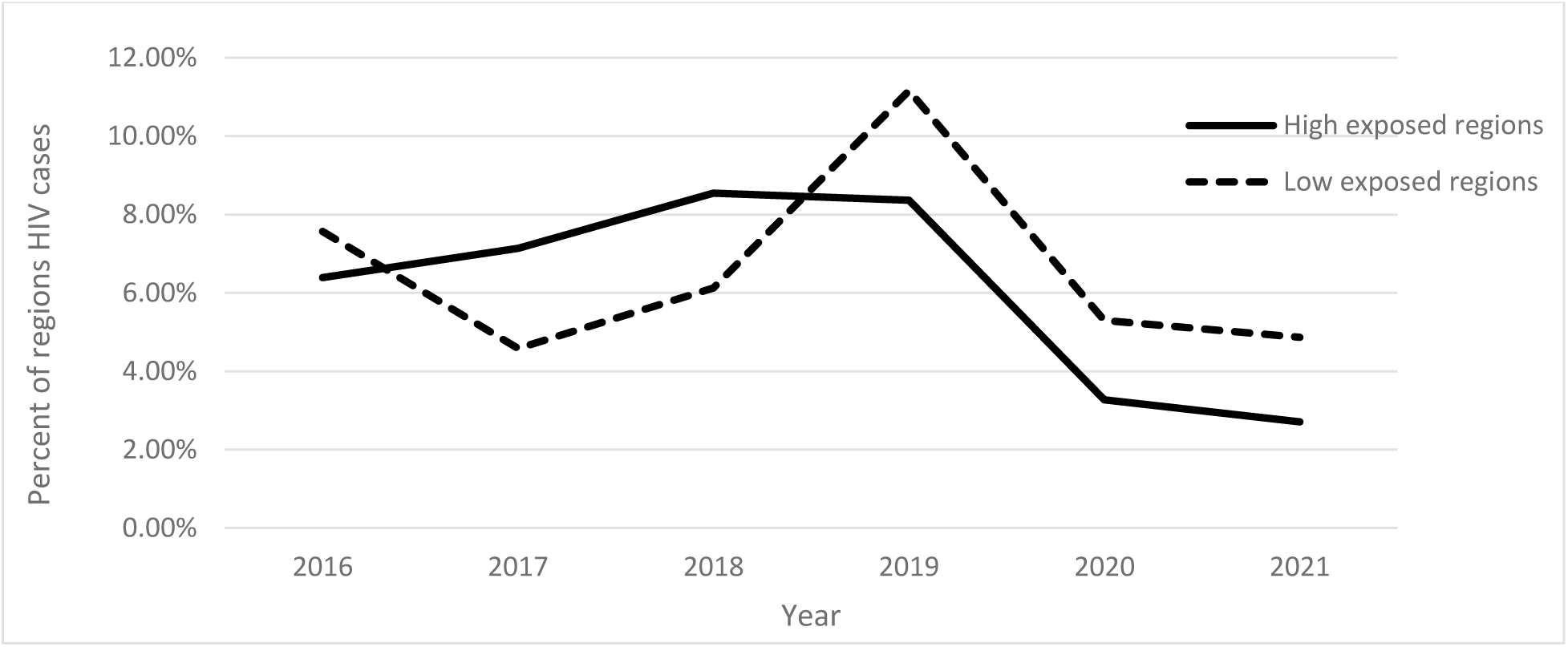
Percent of region’s HIV cases diagnosed at Title X clinics in high vs. low exposure regions from 2016-2021.

### Mixed model results

#### HIV testing at Title X clinics

The null model showed considerable variation in HIV testing between Title X clinics at the regional level (ICC 0.60) indicating that approximately 60% of the total variability in HIV testing is due to differences between regions and justifies the inclusion of region as a random intercept. The regional covariates model showed there was an average decline of 57,346 HIV tests performed at Title X clinics from pre- to post-policy implementation (Table 2). No other covariates were statistically significant. In comparison to the null model, regional covariates significantly improved fit (LRT *X*^2^ (4) = 24.35, P <.001).

**Table 2:**
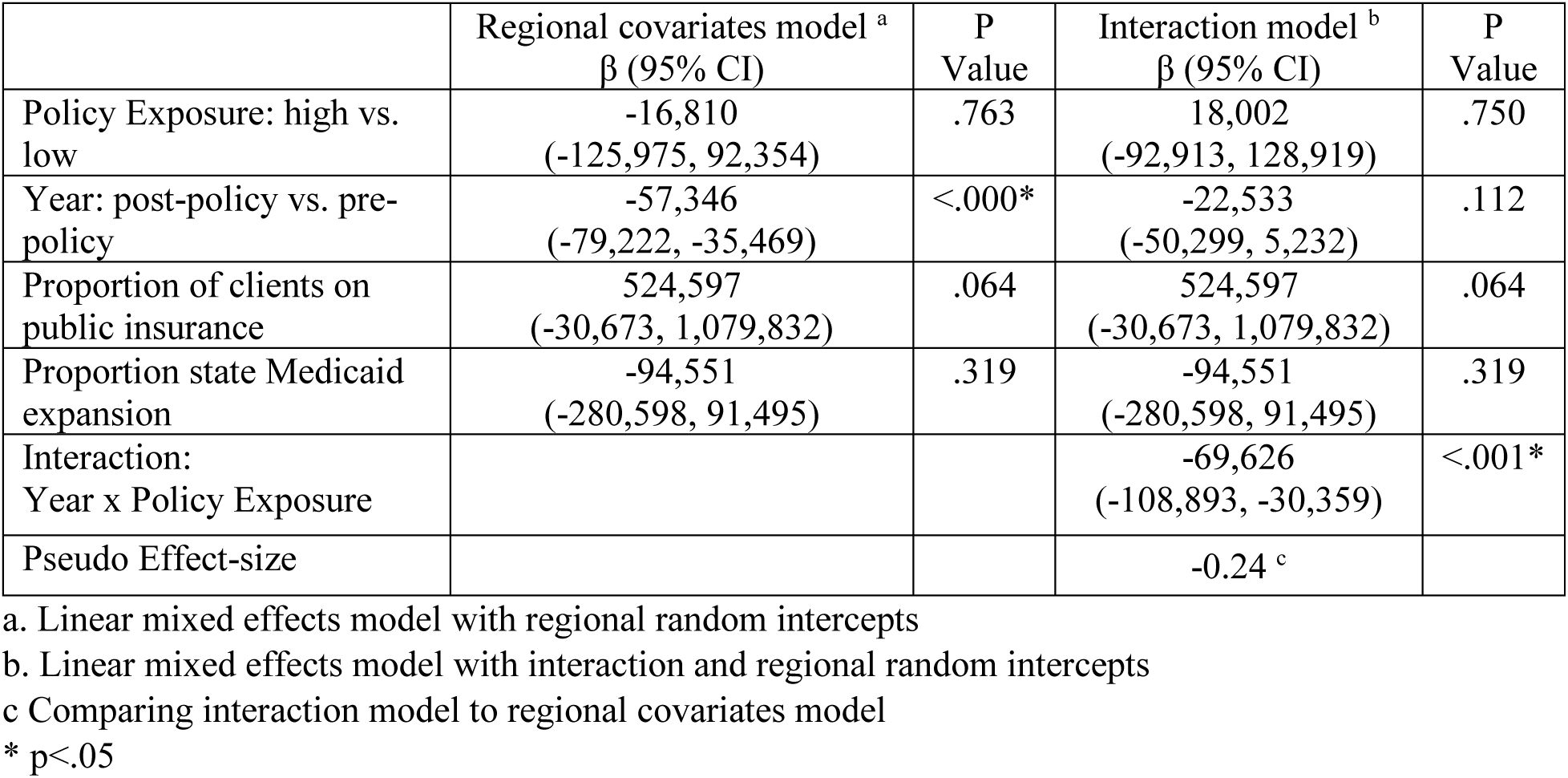
Regional HIV testing at Title X clinics.

|  | Regional covariates model <sup>a</sup><br>β (95% CI) | P<br>Value | Interaction model <sup>b</sup><br>β (95% CI) | P<br>Value |
| --- | --- | --- | --- | --- |
| Policy Exposure: high vs. low | -16,810<br>(-125,975, 92,354) | .763 | 18,002<br>(-92,913, 128,919) | .750 |
| Year: post-policy vs. pre-policy | -57,346<br>(-79,222, -35,469) | <.000* | -22,533<br>(-50,299, 5,232) | .112 |
| Proportion of clients on public insurance | 524,597<br>(-30,673, 1,079,832) | .064 | 524,597<br>(-30,673, 1,079,832) | .064 |
| Proportion state Medicaid expansion | -94,551<br>(-280,598, 91,495) | .319 | -94,551<br>(-280,598, 91,495) | .319 |
| Interaction:<br>Year x Policy Exposure |  |  | -69,626<br>(-108,893, -30,359) | <.001* |
| Pseudo Effect-size |  |  | -0.24 <sup>c</sup> |  |

a. Linear mixed effects model with regional random intercepts
b. Linear mixed effects model with interaction and regional random intercepts
c Comparing interaction model to regional covariates model
\* p<.05

The interaction models showed that, on average, clinics in high exposure regions conducted 69,626 fewer HIV tests compared to clinics in low exposure regions from pre- to post-Policy periods. This difference was statistically significant (95% CI -108,893 to -30,359 P<.001) and improved model fit (LRT *X^2^* (1) = 10.82, P <.001). The pseudo effect size for the interaction term was -0.24, suggesting the Policy had a sizeable negative effect on HIV testing at clinics in high exposure regions.

Results from the sensitivity analyses showed the same trends when the exposure variable was re-coded at 10% and 15% or less net loss of sites. While the interaction term was not significant at 30%, both the exposure and the time variable were significant, likely because of auto-regressive effects or sample size effects. The ordinal coded exposure effects were not significant although the patterns were similar.

#### Proportion of regional HIV cases identified at Title X clinics

The null model showed variation in the proportion of HIV cases diagnosed at Title X clinics between regions (ICC 0.31) and justified the inclusion of random intercepts. The regional covariates model showed Title X clinics in high exposure regions identified, on average, 7% less of the region’s total HIV diagnoses compared to low exposure regions across time (95% CI -10% to -4%; P<.001) (Table 3). The coefficient for state Medicaid expansion was also significant, indicating that regions with higher proportions of states which expanded Medicaid had higher proportions of their region’s HIV cases identified at Title X clinics. The inclusion of regional covariates significantly improved fit (LRT *X^2^* (4) = 15.45, P =.004).

**Table 3:**
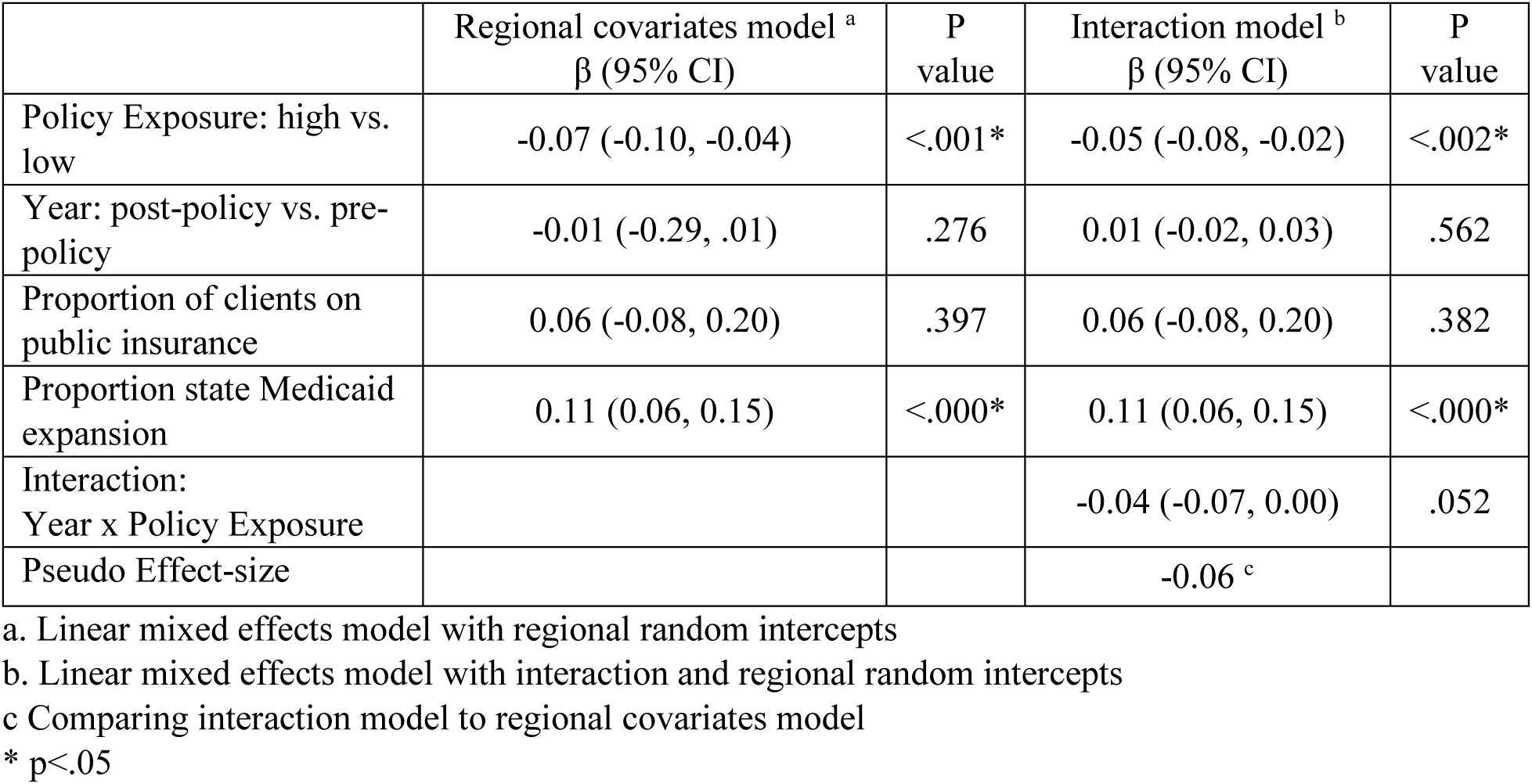
Proportion of region’s total HIV cases identified at Title X clinics.

|  | Regional covariates model <sup>a</sup><br>β (95% CI) | P<br>value | Interaction model <sup>b</sup><br>β (95% CI) | P<br>value |
| --- | --- | --- | --- | --- |
| Policy Exposure: high vs. low | -0.07 (-0.10, -0.04) | <.001* | -0.05 (-0.08, -0.02) | <.002* |
| Year: post-policy vs. pre-policy | -0.01 (-0.29, .01) | .276 | 0.01 (-0.02, 0.03) | .562 |
| Proportion of clients on public insurance | 0.06 (-0.08, 0.20) | .397 | 0.06 (-0.08, 0.20) | .382 |
| Proportion state Medicaid expansion | 0.11 (0.06, 0.15) | <.000* | 0.11 (0.06, 0.15) | <.000* |
| Interaction:<br>Year x Policy Exposure |  |  | -0.04 (-0.07, 0.00) | .052 |
| Pseudo Effect-size |  |  | -0.06 <sup>c</sup> |  |

a. Linear mixed effects model with regional random intercepts
b. Linear mixed effects model with interaction and regional random intercepts
c Comparing interaction model to regional covariates model
\* p<.05

The interaction model showed that on average Title X clinics in high exposure regions identified 4% less of a region’s total HIV diagnoses compared to low exposure regions from pre- to post-Policy (95% CI -7% to -0%; P =.052). A LRT comparing the interaction model to the regional covariates model was non-significant, (χ²(1) = 3.66, P = 0.056). The pseudo effect size for the addition of the interaction term was -0.06, showing a small negative effect of the Policy on the proportion of the region’s HIV diagnoses.

The sensitivity analyses show that when the exposure group threshold is increased to 30% net loss of clinics or using the ordinal exposure variable, that the expansion of Medicaid remains significant, but not at 10% or 15% net loss of sites. While the overall patterns of the estimators remain similar, our exposure variable and interaction term drop below our threshold for significance when the exposure variable is modified.

## DISCUSSION

Our analysis is the first to examine the effect of the 2019 Title X Policy on HIV testing services and diagnoses. We document significant declines in HIV testing and a reduction in the proportion of HIV cases diagnosed at Title X sites as a result of Policy implementation. Our results extend the known negative health impacts of the Policy on contraception provision^21,22^ to include HIV outcomes. Study findings show how policies which target reproductive health providers, such as those in the Title X network, also have implications for HIV service delivery.

The COVID-19 pandemic, beginning in 2020, coincided with Policy implementation and had known effects on HIV testing and diagnoses in the United States.^23^ While there were known difference in COVID-19 effects between urban and rural areas,^24^ we assume relatively comparable effects across the country and our health regions. We find an overall decline in regional HIV testing from pre- to post-Policy periods – likely related to both the Policy and COVID-19. In our interaction models, evidence suggests that the decline in overall regional HIV testing is significantly greater in high exposure regions. This supports the hypothesis that exposure to the Policy is driving the reduction in regional HIV testing at Title X sites beyond trends related to COVID-19.

Results from our sensitivity analyses suggest that the effects of the Policy on declines in HIV testing are robust to other adjustments of our policy exposure, but this trend does not hold for the proportion of HIV diagnoses. This may be an artefact of unbalanced sampling given our small sample since otherwise the patterns and trends remain consistent. Compared to the Policy effect size on HIV tests (-0.24) the Policy effect size for proportion of regional diagnoses was small (- 0.06). This is expected given the multitude of other factors that can influence regional HIV diagnosis rates and testing patterns such as urbanicity, poverty levels, state policy for insurance coverage or protections of sexual minorities etc.^25–27^ In our models we also found that Title X clinics detect a higher proportion of HIV cases in regions with more states that have expanded Medicaid. It is possible that Title X clinics play a more substantial role in HIV diagnoses in states that have expanded Medicaid given Title X’s role in serving clients on public health insurance. The importance of Medicaid in HIV diagnoses was also confirmed by our other models.

While this analysis only examined the impact of the Policy on the Title X program, it is possible that the Policy had broader state or regional impacts while in effect. Evidence suggests that some Title X clinics who left the program and lost federal funding suffered service reductions, staff shortages, or closures due to Policy-related funding cuts.^22,28–31^ These financial difficulties impacted contraception provision^22,31^ and could have led to a reduction or de-prioritization of HIV service delivery. Full clinic closures may have been especially disruptive for a client base which relied on a Title X provider for HIV testing.

The U.S goal to eliminate HIV as a public health threat by 2030^32^ will only be achieved through polices that promote HIV testing and diagnoses. The Title X program is a key contributor to the HIV landscape in the U.S. – diagnosing between 4% and 10% of the total HIV cases between 2016-2021. Strong funding for the Title X program is likely to carry HIV benefits, especially for low-income people who make up 65% of the Title X program’s client base.^33^ Polices that remove restrictions and allow for Title X funds to support the most effective sexual and reproductive health providers will be critical to achieving U.S. HIV goals. Conversely, any future policies akin to the 2019 Policy discussed in the paper will have predictably negative effects for HIV testing and diagnoses rates in the Title X program. U.S. policy makers who aim to support the health of their constituencies must consider this and other data on the negative impacts of restrictions on Title X funding in their decision making.

### Limitations

Outcome data were only available at the regional level, which does not allow for state-level analyses and limits statistical power. While we considered other single-level models using regional indicators or regional clustered standard errors, our mixed model appears the most robust. Despite limited power, statistically significant results were found suggesting there was a significant Policy influence at the regional level. Additionally, no sex-disaggregated Title X data were available for either HIV testing or diagnoses. These data would be important to examine if the Policy had disproportionate effects based on sex. In this non-randomized study design, we examined several potential confounding factors that differed between high and low exposed regions, however other non-measured confounders may exist. Confounding factors could include other state or regional-level policy factors such as abortion legality. Lastly, results of this analysis can only be interpreted as the Policy’s effect on HIV testing and diagnoses in the Title X program, not as the effect of the Policy on the broader HIV landscape in the U.S.

## CONCLUSION

The 2019 Policy on Title X prompted widespread clinic withdrawal and diverted funding away from established HIV testing sites in the U.S. As a result, the program suffered marked declines in HIV testing and diagnoses. Sound health policy that advances efforts to end the U.S. HIV epidemic will support reproductive health clinics, such as those in the Title X program, as key providers of HIV services. Conversely, policies that endanger the Title X network also threaten to weaken the U.S. HIV response. Future discussions on policies governing the Title X family planning program in 2025 and beyond should consider the full consequences for sexual and reproductive health outcomes, including HIV, in the U.S. before implementation.

## Data Availability

All data used in this analysis are publicly available. Collated data may be shared upon request to the corresponding author.

## Authors contribution

The analysis was conceptualized by JS and NR. Data collection was led by DS, JS, and EL and analysis was led by JS and NR. All authors contributed to drafting and editing the final manuscript.

## Declarations

No conflicts to declare. This analysis was not externally funded.

## Appendix 1

Appendix 1.
| Regional designation and changes in Title X site participation |  |  |  |
| --- | --- | --- | --- |
| HHS Region^ | States | Pre- to post-policy net clinic change | Exposure to Policy* |
| 1 | Connecticut, Maine, Massachusetts, New Hampshire, Rhode Island, Vermont | -70% | High |
| 2 | New Jersey, New York, Puerto Rico and the U.S. Virgin Islands | -73% | High |
| 3 | Delaware, District of Columbia, Maryland, Pennsylvania, Virginia, West Virginia | -14% | Low |
| 4 | Alabama, Florida, Georgia, Kentucky, Mississippi, North Carolina, South Carolina, Tennessee | 1% | Low |
| 5 | Illinois, Indiana, Michigan, Minnesota, Ohio, Wisconsin | -39% | High |
| 6 | Arkansas, Louisiana, New Mexico, Oklahoma, Texas | 7% | Low |
| 7 | Iowa, Kansas, Missouri, Nebraska | -9% | Low |
| 8 | Colorado, Montana, North Dakota, South Dakota, Utah, Wyoming | -16% | Low |
| 9 | Arizona, California, Hawaii, Nevada, American Samoa, Commonwealth of the Northern Mariana Islands, Federated States of Micronesia, Guam, Marshall Islands, Republic of Palau | -27% | High |
| 10 | Alaska, Idaho, Oregon, Washington | -67% | High |
| ^ U.S. regions as defined by the Department of Health and Human Services (HHS) |  |  |  |
| *A high exposure designation indicates any region with more than 25% net clinic loss |  |  |  |

## Appendix 2. Model notation and beta-descriptions

Model outcomes include; 1) HIV tests per region (*hiv_test*) and, 2) Proportion of a region’s HIV diagnoses identified at Title X clinics (*prop_pos*).

### Null models

The formulations for these null models were as follows where β0j represents the random intercept for each region and εij is the residual error.

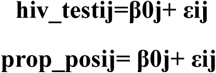

### Regional covariate models

The regional covariates models included covariates for Policy exposure (*exposed*), pre/post Policy time periods (*year_bi*), proportion of Title X clients on public health insurance in the region in 2018 (*public*), and the proportion of states that expanded Medicaid in 2018 (*expand*). The regional covariates models are given by the following where β0j represents the random intercept for each region, β1 represents the average change in the outcome in exposed vs. unexposed regions, β2 represents the average change in the outcome post-policy vs. pre-policy, β3 represents the average change in the outcome per unit increase in the proportion of Title X clients on public health insurance served, β4 represents the average change in the outcome per unit increase in the proportion of states that expanded Medicaid, and εij is the residual error.

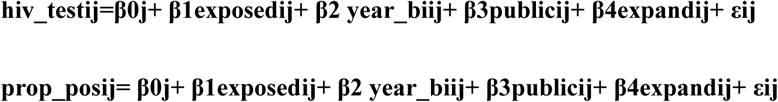

### Interaction models

In the final models, an interaction term was created for exposure and pre/post Policy time (*exposed x year_bi*) and added to the regional covariates models. The interaction models are as follows where β5 is the additive average difference in the outcome for the combined effect of exposed regions and post Policy implementation.

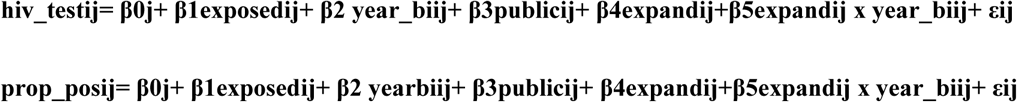

## Notes

### Competing Interest Statement

The authors have declared no competing interest.

### Funding Statement

This study did not receive any funding

